# Communicating with patients and families about difficult matters: A rapid review in the context of the COVID-19 pandemic

**DOI:** 10.1101/2020.04.27.20078048

**Authors:** Stuart Ekberg, Ruth Parry, Victoria Land, Katie Ekberg, Marco Pino, Charles Antaki

## Abstract

**Background:** Pandemics pose significant challenges for healthcare systems, including an increase in difficult discussions about future illness progression and end of life.

**Objectives:** To synthesise existing evidence about communication practices used to discuss difficult matters, including prognosis and end of life, and to use this evidence to make recommendations for clinical practice. The aim of this study was to use rapid review methods to update findings from a previous systematic review published in 2014.

**Data sources:** MEDLINE, EMBASE, CINAHL, PsycINFO, Sociological Abstracts, Web of Science, Scopus, ASSIA and Amed.

**Study eligibility criteria:** Studies using conversation analysis or discourse analysis to examine recordings of actual conversations about difficult matters relating to future illness progression and end of life.

**Study appraisal and synthesis methods:** Data appraisal and extraction procedures used in the 2014 review were modified for this rapid review.

**Results:** Following screening, 18 sources were deemed to meet eligibility criteria, which were added to the 19 sources included in the 2014 systematic review. Synthesis of study findings identified 11 communication practices: providing opportunities for patient or family members to propose matters to discuss (7 out of 37 included sources); seeking a patient or family member’s perspective (6/37); discussing the future indirectly (11/37); discussing the future explicitly (7/37) linking to something previously said or done (11/37); using hypothetical scenarios (13/37); framing a difficult matter as universal (5/37); acknowledging uncertainty (3/37); exploring options (2/37); displaying sensitivity (7/37); emphasising the positive (7/37).

**Limitations:** Dividing work amongst the study authors to enable rapid review may have created inconsistencies.

**Conclusions and implications of key findings:** This synthesis of high-quality evidence from actual clinical practice supports a series of recommendations for communicating about difficult matters during and beyond the COVID-19 pandemic.

## Introduction

From December 2019, the Coronavirus Disease 2019 (COVID-19) rapidly evolved into a pandemic. COVID-19 can cause pneumonia and, in instances where this progresses, can create complications including acute respiratory distress syndrome.^1–3^ For patients requiring intensive care, treatment can include aggressive interventions, such as invasive ventilation.^4–6^

At the time of writing, COVID-19 is posing significant challenges within healthcare systems, including an increase in the frequency of difficult discussions about illness progression and, in some cases, death.^7^ This rapid review synthesises high-quality evidence that can inform how to manage these types of difficult conversations.

In areas where COVID-19 has spread rapidly, healthcare systems have been placed under significant pressure to care for large numbers of patients.^7,8^ Where these systems are placed under extreme strain, clinicians who do not routinely provide critical or end of life care may find themselves needing to discuss difficult matters with patients or their families.^7,9,10^ It is well established that clinicians can feel challenged when they need to discuss prognosis and end of life with patients and their families.^11^ Because communication is an essential clinical skill, considerable scientific evidence has been produced to understand it.^12,13^ There is now a sufficient body of research to inform evidence-based practice when communicating with patients and their families about difficult matters. This review aims to critically review this research and highlight its implications for clinical practice.

### Direct real-life evidence

High-quality evidence about clinical communication is achieved through studies that directly examine video or audio recordings of difficult conversations in real-world clinical practice.^14^ This approach avoids the limitations of self-report methods, which can only provide indirect and partial evidence of what communication is like.^15,16^ Leading approaches to the study of recorded clinical communication are conversation analysis and discourse analysis.^14,17,18^ In contrast to alternative approaches, such as pre-specified coding systems, conversation analysis and discourse analysis employ detailed and inductive methods to understand how specific communication practices function in particular contexts.^17^

Recent decades have seen considerable increase in studies directly examining conversations about difficult matters.^16^ This accumulation of evidence enabled a systematic review, published in 2014, which provides guidance on how to communicate with patients and their families about future illness progression and end of life.^19^ Further growth in evidence has occurred since then.^20^ Given this recent research, and the increased frequency of difficult discussions during the COVID-19 pandemic, the current rapid review updates findings of the 2014 systematic review.

## Methods

### Rapid review approach

This rapid review commenced 30 March 2020 and took four weeks, which is within the typical timeframe for rapid reviews.^21^ The approach was informed by guidelines developed specifically for systematically reviewing and synthesising evidence from conversation analytic and discourse analytic research.^14^

At the time the review was conducted, consensus guidelines for rapid reviews were unavailable.^22,23^ Following suggestions in published research literature,^21,24^ common systematic review methods were adapted for this rapid review. The following adaptations were made: 1) not publishing a protocol before commencing; 2) using rapid review to update a previous systematic review^19^; 3) excluding ‘grey literature’; 4) using only one reviewer to screen search results to identify sources meeting eligibility criteria; 5) not screening the reference lists of included studies to identify additional sources; 6) dividing critical appraisal and data extraction work amongst members of the review team; and 7) having only one reviewer undertake critical appraisal and data extraction from included studies.

### Eligibility criteria

All sources from the 2014 systematic review^19^ were included in the current rapid review. The aim for the rapid review was to add studies published since 2014 that examined audio or audio-visual recordings of actual (i.e. ‘real world’) conversations about difficult matters relating to future illness progression and end of life. As noted, conversation analysis and discourse analysis are leading approaches studying these types of data. The review was therefore restricted to studies employing these approaches. Peer reviewed journal articles and published monographs and book chapters were considered for inclusion. Only studies published in English that examined conversations conducted in English were eligible for inclusion.

### Search strategy

The search strategy employed for the 2014 systematic review^14,19^ was adapted for this rapid review. After initial piloting, one search term (‘future’) was removed to expand the scope of the search and incorporate a greater range of published research. The same databases used in the 2014 systematic review were searched for this rapid review: MEDLINE, EMBASE, CINAHL, PsycINFO, Sociological Abstracts, Web of Science, and Scopus were searched by one reviewer (SE), ASSIA and Amed by another (VL). The search strategy for MEDLINE is available as Supplementary File 1.

Searches were restricted to research published following 1 May 2014, i.e. to dates beyond the previous systematic review.^19^ The final search was conducted on 3 April 2020.

### Study selection

Search results were initially screened by title; where necessary, the abstract or full text were screened to determine whether the study met the eligibility criteria.

### Study appraisal and data extraction

An data extraction form developed by some of the co-authors^14^ was simplified, based on information reported in their 2014 systematic review.^19^ In addition to key information about each study, all fragments of data (i.e. transcripts of real-world conversations) published within the study were extracted. Appraisal and data extraction were conducted simultaneously, to facilitate rapid review. Included studies were divided amongst reviewers (SE, VL, KE, MP, CA) to expedite this process.

### Data synthesis

An aggregative approach, described elsewhere,^25^ was used to compare and connect findings across the included studies. The synthesis was restricted to analytic claims made by the original study authors, rather than those that might be additionally identified by the review team through the pooling of data from across the included studies. Aggregation was undertaken by one reviewer (SE), with critical input from each review team member. Deliberation amongst the team continued until consensus was obtained.

## Results

As shown in Figure 1, 2,382 unique sources were identified through electronic searching of literature published between 1 May 2014 and 3 April 2020. One additional source was identified independently by a review team member. Following screening, 2,365 sources were removed, leaving 18 which met eligibility criteria. These sources were combined with the 19 sources from the earlier systematic review,^19^ resulting in a total of 37 sources for the current review. Further details about the included studies are available at Supplementary File 2.

**Figure 1:**
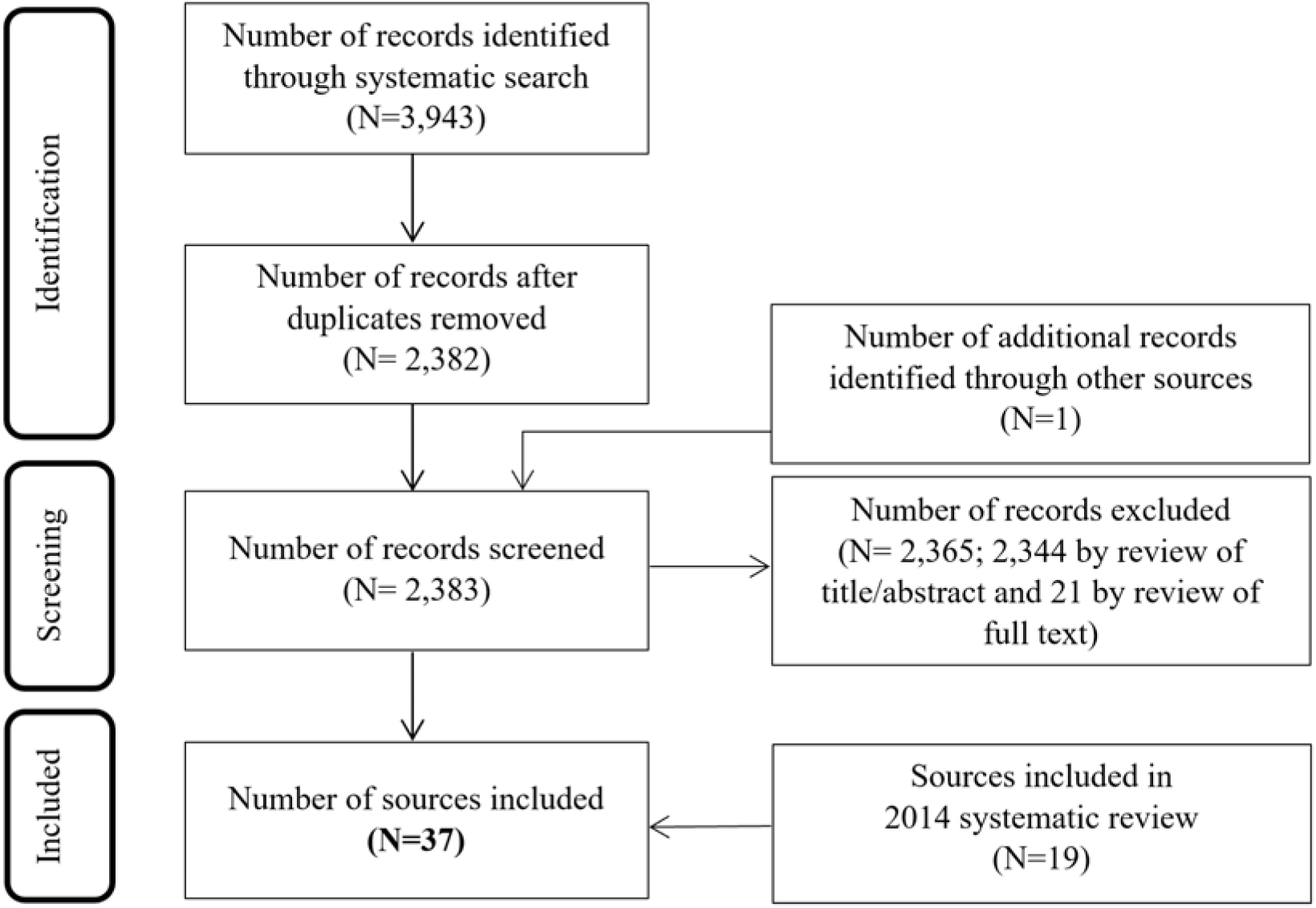
Screening results

Data synthesis identified 11 types of communication practices, which are each described in the below subsections. The support for each communication practice is reported in Table 1.

**Table 1:** Sources supporting each finding

| **Finding** | **Number of sources** | **Settings in which practice has been observed** |
| --- | --- | --- |
| How clinicians provide opportunities for patients or family members to propose matters to discuss | 7 | Counselling<sup>29-31</sup><br>Palliative care/hospice<sup>27,28</sup><br>Oncology<sup>32</sup><br>Psychiatry<sup>33</sup> |
| How clinicians seek a patient or family member's perspective about a specific matter | 6 | Palliative care/hospice<sup>28,34</sup><br>Intensive care<sup>35</sup><br>Oncology<sup>32</sup><br>CALM Therapy<sup>36</sup><br>Psychiatry<sup>33</sup> |
| How clinicians, patients, and family members refer indirectly to the future | 11 | Oncology<sup>32,37-40</sup><br>Palliative care/hospice<sup>34,41,42</sup><br>Primary care<sup>43</sup><br>Internal medicine<sup>44</sup><br>Cardiology<sup>45</sup> |
| How clinicians, patients, and family members refer explicitly to the future | 7 | Oncology<sup>37,38,46,47</sup><br>Palliative care/hospice<sup>34,41,42</sup> |
| How clinicians link to something previously said or done (or not said or done) | 11 | Palliative care/hospice<sup>27,34,48,49</sup><br>Counselling/therapy<sup>29-31,50</sup><br>CALM Therapy<sup>51</sup><br>Psychiatry<sup>33</sup><br>Clinical trial recruitment<sup>52</sup> |
| How clinicians use hypothetical scenarios | 13 | Counselling<sup>29-31,50,53-55</sup><br>Hospice<sup>49</sup><br>Oncology<sup>32</sup><br>Psychiatry<sup>33,56</sup><br>Cardiology<sup>45</sup><br>Primary care<sup>43</sup> |
| How clinicians frame a difficult matter as universal or general | 5 | Oncology<sup>37,38</sup><br>Counselling<sup>30,55</sup><br>Hospice<sup>49</sup> |
| How clinicians acknowledge uncertainty about the future | 3 | Hospice<sup>42</sup><br>Oncology<sup>39</sup><br>Internal medicine<sup>44</sup> |
| How clinicians explore options related to illness progression and end of life | 2 | Oncology<sup>57</sup><br>Neonatal intensive care<sup>58</sup> |
| How clinicians use communication practices that display sensitivity | 7 | Counselling<sup>30,50,55</sup><br>Palliative care/hospice<sup>41,59</sup><br>Oncology<sup>32</sup><br>Clinical trial recruitment<sup>52</sup> |
| How clinicians, patients, and family members emphasise the positive | 7 | Oncology consultations<sup>32,39,60,61</sup><br>Palliative care<sup>27</sup><br>Internal medicine<sup>44</sup><br>Everyday conversations<sup>62</sup> |

### How clinicians provide opportunities for patients or family members to propose matters to discuss

This practice involves clinicians creating opportunities for patients or family members to themselves nominate matters they would like to discuss during a consultation. Often used towards the beginning of consultations when the agenda for the consultation is being set, this practice is also used at other points to ask for additional matters a patient or family member might like to discuss.^26^ A common way of providing such opportunities is through open questions, such as *“Is there anything else you guys wanted to mention or?”*^27^ As identified in the 2014 review, this open-ended design asks about things a patient or family member might like to discuss, but without specifically seeking responses relating to illness progression or end of life.^19^ Although providing these opportunities does not guarantee a patient or family member will raise illness progression or end of life, there are documented instances where patients take the opportunity to raise these matters.^27,28^

### How clinicians seek a patient or family member’s perspective about a specific matter

Clinicians have also been found to seek a patient or family member’s perspective about a more specific matter. These solicitations are often achieved through a ‘perspective display invitation’, which seeks another person’s opinion.^63^ Examples include: *“Do you know her preferences of the kind of quality of life she would want?”*^35^ and *“What do you see as (pause) as the- happening in the future?”*^32^ Patients and family members sometimes responded to these types of questions by raising matters related to illness progression or end of life.^36^ Also, by first soliciting a patient or family member’s perspective, the clinician can then take the patient’s or family member’s perspective into account when providing their own perspective.^34,63^

### How clinicians, patients, and family members refer indirectly to the future

There are a diverse range of practices that can be used to more or less indirectly refer to illness progression or end of life. The most indirect practices included clinicians alluding to the likelihood of illness progression, such as by stating that current treatment has been exhausted*: “I think we’ve gotten as much as we’re going to get from this treatment.”*^40^ The least indirect practices, that nevertheless avoid explicit references such as ‘death’ or ‘dying’ are euphemisms such as *“when he passes.”*^41^ Towards the middle of this spectrum of indirect practices are references to time, such as *“I think he probably has a limited amount of time now.”*^42^ There is some evidence that indirect discussions about illness progression and end of life is the default way of talking about such matters. This is particularly the case when the person being discussed is either involved in the conversation or is a significant for one or more parties to the conversation.^28,32,34,41^

### How clinicians, patients, and family members refer explicitly to the future

In contrast to practices that discuss future deterioration and end of life indirectly, these studies explore instances where such matters are made explicit. On some occasions, discussions about end of life were initiated by clinicians indirectly, and subsequently made explicit by patients.^32,34^ In general, clinicians’ explicit references to end of life appear to be more common after the matter has been made explicit by patients themselves.^37,38^

There are, however, exceptions where clinicians initiate more explicit discussions about illness progression and end of life. Sometimes, in instances where patients or family members seem to avoid talking about illness progression or end of life, clinicians can respond by referring to these matters more explicitly,^32,34^ such as with: *“Do you worry about what’s coming?”*^34^ Another exception is when clinicians explicitly discuss death to promote acceptance of treatment. The following is one such instance: *“If we don’t do it…it will come back and if it comes back it will be deadly.”*^46^ Evidence suggests these explicit references to death to warrant acceptance of treatment only tend to occur after patients have displayed some resistance to treatment recommendations.^46,47^

### How clinicians linked to something previously said or done (or not said or done)

Studies found that clinicians can mention something said or done in the recent or distant past that is related to illness progression or end of life, then use this to promote further discussion about these matters. Examples include: *“So coming back to what you were saying before…part of it is the fear of what might happen?”*^34^; *“Do you remember when you first came on the ward here?…Things were pretty desperate…And we got you on a little syringe pump with the pain medicine in?”*^49^ There are documented instances where patients respond to such solicitations with matters relating to illness progression or end of life.^34^ Clinicians can also link to something a patient has not said, to provide a basis for asking about it: *“You haven’t mentioned AIDS as a concern today. How much of a concern is that?”*^31^

### How clinicians use hypothetical scenarios

Hypothetical future scenarios can be used to foster discussion about matters relating to illness progression or end of life.^19^ Examples include: *“And if there was a bit of uh bang, if there was a bit’v bleeding or some other crisis, how would you want to handle that do you think?”*^49^; *“If you- supposing- I mean this is just supposing, supposing you had got infected or were to get infected…”*^30^ There are contexts where these practices appear to be particularly effective at occasioning discussion about illness progression or end of life. These contexts include circumstances where a patient or family member has displayed reticence to discuss these matters, or to question a patient or family member’s expressed plans or expectations for the future.^19,49^

### How clinicians frame a difficult matter as universal or general

In contrast to hypothetical scenarios, which involve discussions related to the individual patient, this type of practice involved framing matters abstractly, as something that could be universally faced by anyone rather than a particular patient specifically.^19^ This abstract framing occurs in the following instance: *“…sometimes when people are really unwell…what we do is we get them some medicine at home.”*^49^ There is some evidence that universal statements are more likely in relation to matters that have not been raised by a patient or family member in the past.^30^ Their use softens the direct relevance of the matter being discussed to the patient.^49^

### How clinicians acknowledge uncertainty about the future

This practice involves clinicians using expressions that qualified their level of certainty, as well as explicit statements of uncertainty. The first part of the following instance includes qualifying expressions (‘looks like’ and ‘probably’), and the second part an explicit statement of uncertainty: *“This looks like the last days probably…We have learned that we have no idea to predict how many.”*^42^ This practice demonstrates that prognostic uncertainty does not need to prevent discussions about difficult matters such as illness progression and end of life.^42^

### How clinicians explore options related to illness progression and end of life

By exploring options, clinicians can make apparent to patients and family members different possibilities for future care. Sometimes clinicians present options as equivalent, while other times they convey a preference for one option. The studies included in this review span this diversity, with one study finding that exploring options informed collaborative decision making in the context of illness progression,^58^ the other finding that exploring options was used to negatively appraise particular treatment options in the context of illness progression.^57^ Due to the small number of included studies, and their distinct findings, further research is warranted to understand broader implications for clinical practice.

### How clinicians use communication practices that display sensitivity

Additionally to practices described above, such as discussing illness progression and end of life indirectly, there are other practices that clinicians can empathise with a patient or family member’s situation, during discussions of illness progression or end of life. Communication practices that displayed sensitivity include explicit forms of sensitivity, such as displays of empathy that show a clinician’s understanding of a patient’s emotional experience: *“I know it’s not always the easiest thing to uh to chat about.”*^59^ Evidence also suggests silence or brief responses such as ‘mm’ can be effective once talk about difficult matters has been broached.^52,55^ Several of the included studies suggested that talk about difficult matters is more likely to contain hesitations, delays, cut-off words, and repeated words or phrases.^41,50^ As noted in the 2014 review,^19^ there are only some limited observations that consider how non-verbal behaviour, such as touch, can be used to convey sensitivity.

### How clinicians, patients, and family members emphasise the positive

These studies showed that as discussions about difficult matters progress, this often involved shifts away from discussing these difficulties to talking about something more positive, as in the following: *“And I will tell you this is frankly a bit on the outer edge of our ability to get rid of. I want to be very candid with you. But I do believe that we can do this.”*^39^ As noted in the 2014 review, such practices can be used to sustain hope and preserve relationships, but they can also divert the conversation, thereby preventing further talk about difficult matters.^19^

## Discussion

This rapid review has synthesised findings from studies examining difficult conversations about illness progression and end of life. The review focuses on high-quality evidence delivered by studies examining real-world conversations to understand how specific communication practices function in particular contexts.^14,17,18^ Since publication of a systematic review on this topic in 2014,^19^ evidence in this area has almost doubled. As reported in Box 1, the synthesis of findings from these studies has enabled the generation of 10 recommendations for clinicians who find themselves needing to discuss difficult matters about illness progression or end of life with patients or their families.

#### Box 1: Summary of evidence-based guidance

##### Try to find out what a patient or family member would like to get out of a conversation

Where possible, create opportunities for patients or family members to nominate matters they would like to discuss. They may indicate their interest in discussing illness progression or end of life. If they do not, it may nonetheless be possible to get a sense of how open, or reluctant, they might be to engage with such matters. What you say next can be informed by this.

##### Try to find out a patient or family member’s perspective about the future

Before offering your own perspective about a patient’s future, try to understand a patient’s or family member’s perspective about this matter. This will help you to take that perspective into account when deciding how to offer your own perspective and to choose how strongly to push patients and family members to engage with your perspective.

##### If a patient or family member talk about the future indirectly or allusively, try to do the same

In many societies, it is common for dying and death to be discussed indirectly. If patients or family members talk about the future indirectly and this does not appear to create the possibility for misunderstanding, try to use similar language. Over time, they may come to discuss the future more directly, in which case you can adjust your language accordingly.

##### Sometimes the future may need to be talked about more directly

Sometimes patients or family members are willing to talk openly about illness progression or end of life – either from the outset of a conversation, or increasingly as the conversation progresses. If you think it is important to discuss these matters explicitly, and a patient or family member has not displayed a willingness to do so, or has resisted indirect attempts to do so, consider how you might broach this sensitively. Some of the other strategies listed below may be useful.

##### Connect what a patient or family member has said to what you’re saying now

To promote further talk about future illness progression or end of life, try bringing up something the patient or family member has mentioned before about the future, then use this to promote further discussion about this matter. You can help them link concerns they have already expressed, with concerns about and plans for end of life.

##### Consider using hypothetical scenarios

Talking about the future hypothetically means patients and family members do not need to agree that this is necessarily how their future will transpire. Evidence suggests people can be more open to engaging in these types of hypothetical discussions.

##### Where appropriate, frame difficult matters as universal

If you need to mention a difficult matter, but are unsure how a patient or family member will react, consider framing this matter, where possible, as something that applies universally (e.g., “when people are very ill…”). This practice can be useful when you want to raise something that a patient or family member hasn’t already hinted at, or where you want to provide them with an opportunity to recognise its relevance to them, without forcing them to do so.

##### Be clear about uncertainty

Even when illness progression and end of life are certain, explain things that are less certain, such as the timeframe for progression.

##### Communicate to display sensitivity

There are many ways you might display sensitivity when discussing difficult matters relating to illness progression and end of life. In addition to empathic statements, consider additional practices such as allowing periods of silence.

##### Acknowledge positives, but not too soon

Emphasising positive things, such as what can be done for a patient, can be useful for sustaining hope. When people are discussing difficult matters, however, starting to talk about positive things can end the difficult discussion. For this reason, consider when is the right time to move from the difficult aspects of a discussion, to the more positive dimensions.

The evidence-based recommendations listed in Box 1 are not prescriptive, nor do they recommend clinicians use scripted phrases. This approach recognises that the contingencies of communication mean these social encounters can rarely – if ever – ‘follow the script.’^64^ The recommendations reflect this complexity, to help explain, for instance, why people discuss sensitive future matters indirectly in some circumstances and explicitly in other circumstances. The communication practices described in Box 1 range from some that are relatively more cautious and indirect, to those that are relatively more forceful and direct. It is important to consider, on a case-by-case basis, which approaches are likely to be most suitable. As always in evidence-based practice, quality evidence should inform, but not replace, clinicians’ decisions about how to provide care that is appropriate for individual patients and their circumstances.^65^

There are already many strategies and frameworks designed to inform the conduct of discussions about difficult matters related to illness progression and end of life.^66,67^ Prominent contemporary approaches include the SPIKES protocol,^68^ VitalTalk,^69^ and the Serious Illness Conversation Guide (SICG).^70^ The recommendations made in Box 1 have important similarities and differences to these resources. For instance, recommendations to elicit the patient’s perspective (SPIKES), assess perception of illness (VitalTalk), and assess illness understanding (SICG) are consistent with the second recommendation listed in Box 1. This review also documents practices that extend beyond those recommended in these available resources, such as considering why specific communication practices such as communicating indirectly and using hypothetical scenarios may be useful. This highlights a key advantage of using direct empirical evidence to understand what constitutes effective communication in clinical settings.^71^

The studies included in this rapid review examine data collected before the COVID-19 pandemic. Although they do not directly examine difficult conversations in the context of a pandemic, the included studies consider a diverse range of conversations collected across many different clinical specialties. The identification of common types of communicative practices used across these diverse settings increases confidence that the findings of this review are transferrable to the difficult discussions clinicians must have with patients and family members during situations like the COVID-19 pandemic. The broad focus of this review means its findings can also be transferred to other difficult conversations about illness progression and end of life that occur in clinical settings outside of pandemics.

The rapid review methodology adopted for this study means there may be limitations to its findings. For instance, dividing work on the quality assessment and data extraction phases of the review amongst the review team may have introduced inconsistencies in work. In particular, this could have inhibited scope to identify and document similar findings across studies, thus preventing these being incorporated within the subsequent data synthesis phase. For this reason, this review updates but does not replace a systematic review conducted in 2014,^19^ which had greater scope to systematically review and synthesise findings than was likely to have been the case in the current study.

Considerable progress has been made in developing high-quality evidence to inform difficult conversations about illness progression and end of life. Research based on direct and detailed analysis of real life difficult discussions that have been audio- or video-recorded has almost doubled since a systematic review was published in 2014.^19^ Nevertheless, further research is likely to yield additional insights into the nature of these conversations. In particular, further research is needed to understand ways clinicians acknowledge uncertainty and explore options with patients and their families. Additional research is also needed to understand how clinicians manage conversations about difficult matters when healthcare systems are placed under considerable strain, such as during the COVID-19 pandemic. Through detailed analysis of the difficult conversations that occur in healthcare, an increasingly clearer understanding will emerge to enhance this fundamental part of clinical practice, and to thereby improve the experience of patients and their families.

### Contributor information

This rapid review updates a previous systematic review conducted by RP and VL. SE and RP conceived and designed the current rapid review. SE and VL conducted the literature search and screened articles for inclusion in the review. SE, VL, KE, MP, and CA conducted study appraisal and data extraction. SE conducted the synthesis, with critical feedback from all authors. SE led the initial drafting of the manuscript. All authors critically reviewed and contributed to re-drafting of the manuscript. SE is the guarantor of the research reported here.

### Patient and public involvement

Due to time constraints, this rapid review was undertaken without patient or public involvement.

## Data Availability

This article synthesises research findings that have already been published.

